# Large-scale Analyses of CAV1 and CAV2 Suggest Their Expression is Higher in Post-mortem ALS Brain Tissue and Affects Survival

**DOI:** 10.1101/2022.11.04.22281798

**Authors:** Brett N Adey, Johnathan Cooper-Knock, Ahmad Al Khleifat, Isabella Fogh, Philip van Damme, Philippe Corcia, Philippe Couratier, Orla Hardiman, Russell McLaughlin, Marc Gotkine, Vivian Drory, Vincenzo Silani, Nicola Ticozzi, Jan H. Veldink, Leonard H. van den Berg, Mamede de Carvalho, Susana Pinto, Jesus S. Mora Pardina, Monica Povedano, Peter M. Andersen, Markus Weber, Nazli A. Başak, Christopher E Shaw, Pamela J. Shaw, Karen E. Morrison, John E. Landers, Jonathan D. Glass, Patrick Vourc’h, Richard JB Dobson, Gerome Breen, Ammar Al-Chalabi, Ashley R Jones, Alfredo Iacoangeli

## Abstract

Caveolin-1 and Caveolin-2 (CAV1 and CAV2) are proteins associated with intercellular neurotrophic signalling. There is converging evidence that CAV1 and CAV2 (CAV1/2) genes have a role in ALS. Disease-associated variants have been identified within CAV1/2 enhancers, which reduce gene expression and lead to disruption of membrane lipid rafts. Using large ALS whole-genome sequencing and *post-mortem* RNA sequencing datasets (5987 and 365 tissue samples, respectively), and iPSC-derived motor neurons from 55 individuals, we investigated the role of CAV1/2 expression and enhancer variants in the ALS phenotype. We report a differential expression analysis between ALS cases and controls for CAV1 and CAV2 genes across various *post-mortem* brain tissues and three independent datasets. CAV1 and CAV2 expression was consistently higher in ALS patients compared to controls, with significant results across the primary motor cortex, lateral motor cortex, and cerebellum. We also identify increased survival among carriers of CAV1/2 enhancer mutations compared to non-carriers within Project MinE and slower progression as measured by the ALSFRS. Carriers showed a median increase in survival of 345 days. These results add to an increasing body of evidence linking CAV1 and CAV2 genes to ALS. We propose that carriers of CAV1/2 enhancer mutations may be conceptualised as an ALS subtype who present a less severe ALS phenotype with a longer survival duration and slower progression. Upregulation of CAV1/2 genes in ALS cases may indicate a causal pathway or a compensatory mechanism. Given prior research supporting the beneficial role of CAV1/2 expression in ALS patients, we consider a compensatory mechanism to better fit the available evidence, although further investigation into the biological pathways associated with CAV1/2 is needed to support this conclusion.

## Introduction

Amyotrophic lateral sclerosis (ALS) is a fatal neurodegenerative disease affecting upper and lower motor neurons. It is characterised by the progressive loss of motor function, leading to muscle weakness, difficulty breathing and swallowing, and paralysis. There is currently no treatment, with a mean life expectancy of 3 years (1). ALS is comorbid with fronto-temporal dementia (FTD), with an estimated 50% of ALS patients experiencing impaired executive function (2,3). These diseases are often conceptualised as two ends of a disease spectrum with a shared pathogenesis and clinical overlap (4,5).

Individuals who have a first degree relative with ALS are twice as likely than average to develop ALS (6), and patients with a family history (familial ALS) make up approximately 5-10% of cases (7). A pathogenic variant for familial patients can be identified in over 50% of cases (8). However, most cases have no family history (sporadic ALS), and the majority have no identified genetic aetiology. A recent genome-wide association study (GWAS) estimates the narrow-sense heritability of ALS due to SNPs at 8.5% (9). This represents a minimum heritability value based upon variation of SNPs included in sequencing arrays. Broad sense heritability estimations for ALS vary between 43-53% (10,11). Of all currently known pathogenic variants, the most common is a hexanucleotide repeat expansion within the *C9orf72* gene, which accounts for ∼30-40% of familial cases and 5-10% of sporadic cases (12,13). Individuals with this mutation display an earlier age of onset and faster disease progression (14). A further ∼5% of sporadic cases are attributable to mutations in *SOD1, FUS*, and *TARDBP* genes (15).

Despite these known genetic variants, a large proportion of ALS heritability remains unaccounted for. Most ALS genetic studies focus on the study of rare single nucleotide variants (SNVs) and small insertions and deletions (indels) in the coding regions of the genome, or on common single nucleotide polymorphisms (SNPs). As a consequence, structural and rare variants in non-coding regions of the genome are largely under-investigated and could represent a potential source of the missing heritability (16–18).

Caveolin-1 and Caveolin-2 (CAV1 and CAV2, or CAV1/2) genes code for proteins that are associated with the function of membrane lipid rafts. These are regions of low fluidity within the cellular membrane, which act as anchoring points for intercellular signalling (19). Converging evidence links CAV1 and CAV2 genes to ALS pathology; CAV1 is associated with neuronal survival and is upregulated during induced ischemia in mice, aiding uptake of extracellular vesicles and reducing apoptosis (20). CAV1 may also play a role in the cognitive decline associated with ALS/FTD (21), with overexpression increasing neuroplasticity, pro-growth signalling, learning, and memory in mice (22,23). Additional evidence using male *SOD1* mice showed that promotion of neuron-specific CAV1 expression increases body weight and improves longevity and motor function (24). In a subsequent mouse study, subpial administration of synapsin-promoted CAV1 also increased survival, although saw no changes to body weight or motor function (25). Conversely, increased neurodegeneration and synaptic reduction was observed in CAV1 knock-out mice (26).

In humans, CAV1 coding regions are enriched for ALS-associated variants and CAV1 and CAV2 enhancer mutations are significantly associated with increased risk of ALS (17). An expression analysis revealed that two mutations within CAV1 and CAV2 enhancer regions reduced CAV1/2 expression in patient-derived non-neuronal cells, which was supported by CRISPR-Cas9 editing in neuronal cells (17). Together, evidence from human and mouse studies indicate that CAV1/2 is neuroprotective, and CAV1/2 mutations are a risk factor for ALS pathology, likely as a consequence of reduced gene expression.

In this study we aim to investigate whether these mutations lead to differences in disease-related phenotypes, as well as changes in ALS risk, and explore whether CAV1/2 expression plays a role in the disease beyond enhancer mutations. In the first set of analyses, we used an RNA-sequencing pipeline to perform expression analysis of the CAV1 and CAV2 genes. The results supported our hypothesis that CAV1 and CAV2 genes would be differentially expressed between ALS cases and controls, with patients showing increased expression. In the second set of analyses, we investigated differences in survival duration and age of onset between ALS patients with and without CAV1/2 enhancer mutations. Considering evidence that CAV1/2 enhancer mutations reduce CAV1/2 expression, and that CAV1/2 expression is beneficial to ALS phenotypes, we hypothesised a reduced survival duration and earlier age of onset in ALS patients who have CAV1 or CAV2 enhancer mutations. Results were opposite to our expectation, showing increased survival duration among carriers of CAV1/2 enhancer mutations. No difference in age of onset was observed between groups.

To confirm whether differential expression of CAV1/2 occurred in neurons specifically, we ran an RNA-seq expression analysis in iPSC-derived motor neurons (MNs) from ALS patients and neurologically normal controls. Additionally, we examined the presence of correlation between expression of CAV1/2 in the iPSC-derived MNs and survival, age of onset, and disease progression as measured by the ALSFRS.

## Methods

### Sequencing and clinical data

#### Datasets for RNA-seq differential expression

RNA-seq datasets for the differential expression analyses were obtained from TargetALS at the New York Genome Centre (NYGC) (NCBI GEO ID: GSE116622 & GSE124439), the Florida Mayo Clinic (NCBI GEO ID: GSE67196), and the King’s College London and MRC London Neurodegenerative Diseases Brain Bank (15,27,28).

Sample collection and data generation were previously described (15). Briefly, frozen human *post-mortem* samples were used in all cases, and tissue was taken across multiple brain areas. The KCL MRC Brain Bank samples were taken from the primary motor cortex. The Mayo Clinic samples were obtained from the lateral hemisphere of the cerebellum, Brodmann area 9 and 44 (prefrontal cortex) and Brodmann area 4 (primary motor cortex). The Target ALS (NYGC) samples were obtained from the cerebellum, the lateral and medial motor cortex, and various locations within the frontal cortex.

#### Project MinE

Whole genome sequencing and clinical data of ALS cases from Project MinE (data freeze 2) were used for the survival and age of onset analyses (29,30). Samples were filtered to remove common variants (MAF > 0.01) in the enhancer regions of CAV1 and CAV2 genes, which are defined in Cooper-Knock et al., 2020 (17). Individuals with missing data for sex, survival, and age of onset for the corresponding analysis, or those that failed quality controls (30) were removed. This retained 5987 cases for analysis, including 44 individuals with at least one CAV1 or CAV2 enhancer mutation. Data generation and whole-genome sequencing quality controls, including principal component analysis, were previously described (29–31).

#### AnswerALS

Total RNA-seq gene expression profiling of iPSC-derived MNs and phenotype data were obtained for 55 ALS patients and 15 controls from AnswerALS (32). Gene expression was normalized for gene length and then sequencing depth to produce transcripts per kilobase million (TPM). Age of onset and disease status were available for all individuals and these parameters were used to check for the correlation between expression of top-ranked RefMap ALS genes and age at disease onset.

### Data Analysis

#### RNA-seq Differential Expression Analysis

An RNA-seq based differential expression analysis was performed for CAV1 and CAV2 genes on samples across three datasets. Detailed protocol of library preparation is described by Tam et al. 2019 for TargetALS samples (33), Prudencio et al. 2015 for Mayo Clinic samples (34), and Jones et al. 2021 for the KCL MRC Brain Bank samples (15). *Figure 1* illustrates the stages performed in the RNA-seq analysis.

**Figure 1.**
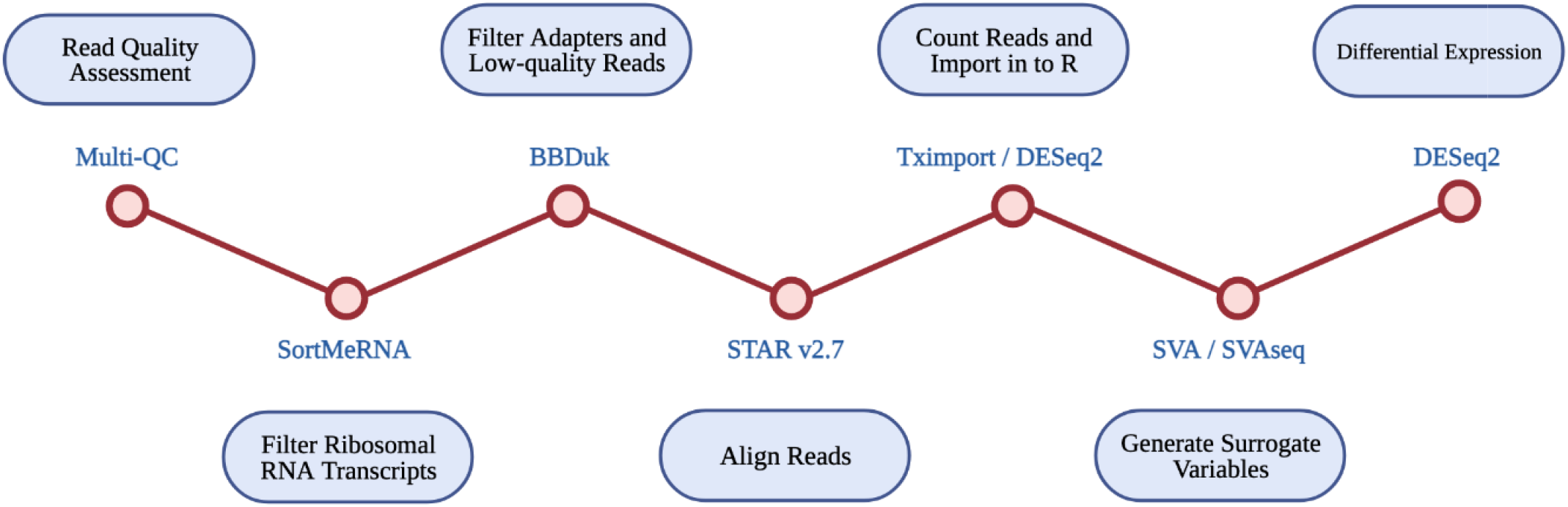
Diagrammatic representation of RNA-seq differential expression pipeline. Each RNA-seq step is shown in the blue circles, with the tool used at each step given beside each red circle.

Multi-QC was used for all datasets to assess read quality pre- and post-alignment. The removal of ribosomal RNA transcripts was achieved by filtering with SortMeRNA, using rRNA databases. BBDuk was used to filter adapters and low-quality reads. RNA-seq reads were aligned with STAR v2.7 using the GRCh37.89 reference genome.

Read counts were imported into R using Tximport and DESeq2. Only transcripts with at least 10 reads were retained for analysis. Available data for disease status, gender, quintiles of age, quintiles of PMI, RIN, and flow-cell were imported into R. SVA and SVAseq were used to generate surrogate variables for each sample, which estimate expression heterogeneity. These were included as covariates in subsequent analyses to control for unaccounted confounding factors such as cell heterogeneity and extraneous variation.

Raw read counts were supplied to DESeq2, which was used to perform a differential expression analysis across ALS cases and controls. Differential expression was estimated using log_2_ fold-change, a wald test, and FDR *p*-value correction. Analyses were run using covariates of age, gender, *post-mortem* delay, RIN, and surrogate variables, where data was available.

The final differential expression results were meta-analysed for each brain tissue type using the Stouffer method (35). This uses the p-value, sample size, and log_2_ fold-change from each dataset to produce meta-analysed test statistics, and considers the direction of effect.

#### Project MinE Survival and Age of Onset Analyses

Multiple cox proportional hazard survival analyses were run and visualised in R using the *survival* and *survminer* packages. These analyses were to assess whether the presence of CAV1/2 enhancer mutations impacts patient survival. Analyses were run with sex at birth and age of onset as covariates, using individuals with no CAV1/2 mutations together with: CAV1 mutations only, CAV2 mutations only, and individuals with mutations in either gene.

C9-related ALS is characterised by different clinical presentation (36,37), earlier age of onset, and faster disease progression compared to non-C9 ALS, suggesting a separate disease mechanism (14). Analyses were therefore run with and without individuals carrying a pathogenic repeat expansion of the *C9orf72* gene (14) to assess whether increasing sample homogeneity would reveal a stronger effect of CAV1/2 mutations on survival. Analyses were additionally run excluding samples from patients with other well-known ALS mutations (*SOD1, FUS, TARDBP*), and matching samples based on nationality. Finally, survival analyses were run when stratifying samples by type of CAV enhancer mutation (CAV1 or CAV2).

A second set of analyses were run to determine whether CAV1/2 status affected age of onset, using sex at birth as a covariate. These were a linear regression and cox proportional hazard models, run in R using the survival package. Analyses were run with and without carriers of a pathogenic *C9orf72* repeat expansion. They compared samples with no CAV1/2 mutation to: [1] samples with CAV1 enhancer mutations, [2] samples with CAV2 enhancer mutations, and [3] samples with either mutation.

## Results

### Samples and datasets

#### Differential Expression Analysis Datasets

Samples were matched across disease status by age and sex within each dataset, where data permitted. Cases were comprised of samples from sporadic and familial ALS patients, including *C9orf72-* and *SOD1*-associated ALS. Control samples were obtained from individuals with non-neurological or non-ALS disease. An outline of each dataset is provided in *Figure 2*.

**Figure 2.**
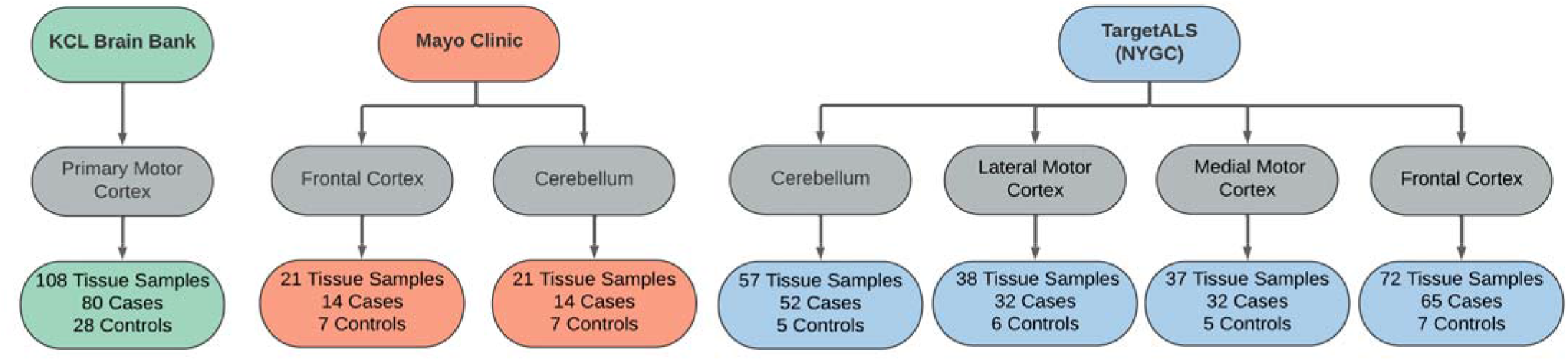
Sample overview across the RNAseq datasets used in the differential expression analyses. Datasets were obtained from the KCL Brain Bank (green), Mayo Clinic (orange), and TargetALS (NYGC; blue).

#### Project MinE dataset for CAV1/2 enhancer mutation Analyses

CAV1/2 enhancer variants of MAF < 0.01 in gnomAD were filtered prior to analysis. 5,987 samples passed the quality controls and were used for analysis. Of these, 356 were carriers of the *C9orf72* repeat expansion. In total, 44 patients had at least one CAV1/2 enhancer mutation, of which, 34 were carriers of CAV1 mutations, and 10 were carriers of CAV2 mutations. *Figure 3* shows sample sizes for the four primary Project MinE survival analyses.

**Figure 3.**
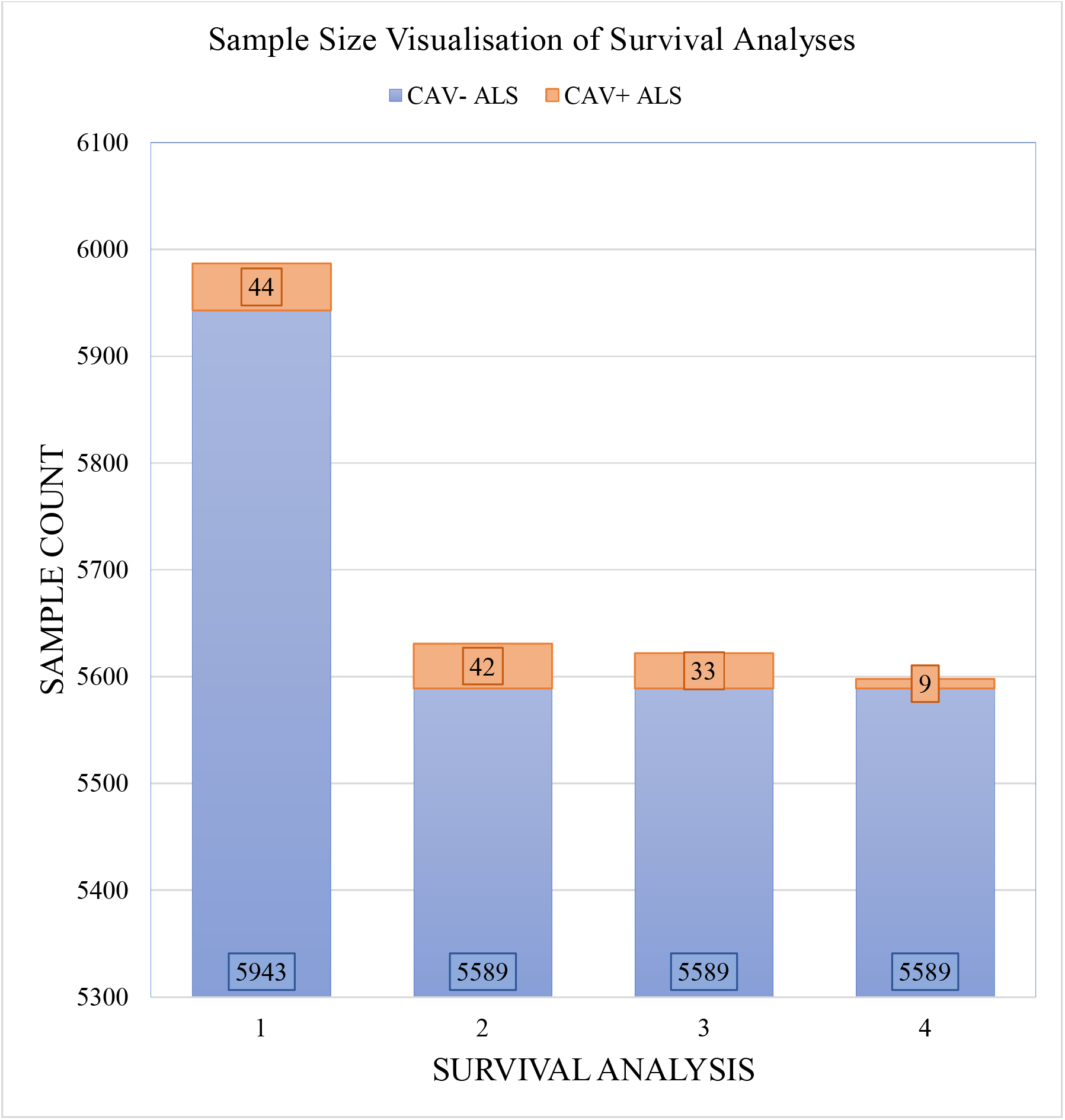
Sample sizes for each Project MinE survival analysis. Samples are divided by those with CAV1/2 enhancer mutations (orange) and without (blue). Analyses are: [1] Full dataset including all rare mutations [2] excluding C9orf72 samples [3] with CAV1 and CAV-samples only [4] with CAV2 and CAV-samples only. CAV-refers to patients who do not carry CAV1/2 enhancer mutations and CAV+ refers to those who do.

### Bulk RNAseq reveals higher expression of CAV1 and CAV2 in ALS patient tissue compared to controls

Considering converging evidence that CAV1/2 genes are neuroprotective and the previous association between ALS disease status and CAV1/2 enhancer regions, we hypothesised that CAV1 and CAV2 genes would be differentially expressed between ALS patients and controls within brain tissue. Results from the differential expression analysis for CAV1 and CAV2 are outlined in *Table 1* and shown in violin plots in *figure 4*. CAV1 showed statistically significant differential gene expression within the KCL primary motor cortex (Log2FC = 0.396, *p* = 0.04) and the NYGC cerebellum (Log2FC = 0.751, *p* = 0.02). CAV2 was differentially expressed in the primary motor cortex within the KCL BrainBank sample (Log2FC = 0.183, *p* = 0.01), in addition to the cerebellum (Log2FC = 0.669, *p* = 0.004) and lateral motor cortex (Log2FC = 0.691, *p* = 0.029) within Target ALS (NYGC) samples. Dataset-tissues almost universally showed a positive log_2_ fold-change (with the exception of the NYGC frontal cortex), suggesting that CAV1/2 is consistently upregulated among ALS cases. This direction of effect is contrary to previous evidence if we conclude that a higher expression level in cases corresponds to gene expression increasing ALS risk. However, this aligns with a compensatory model, in which expression of CAV1/2 genes is increased to mitigate ALS-related pathology.

**Table 1.**
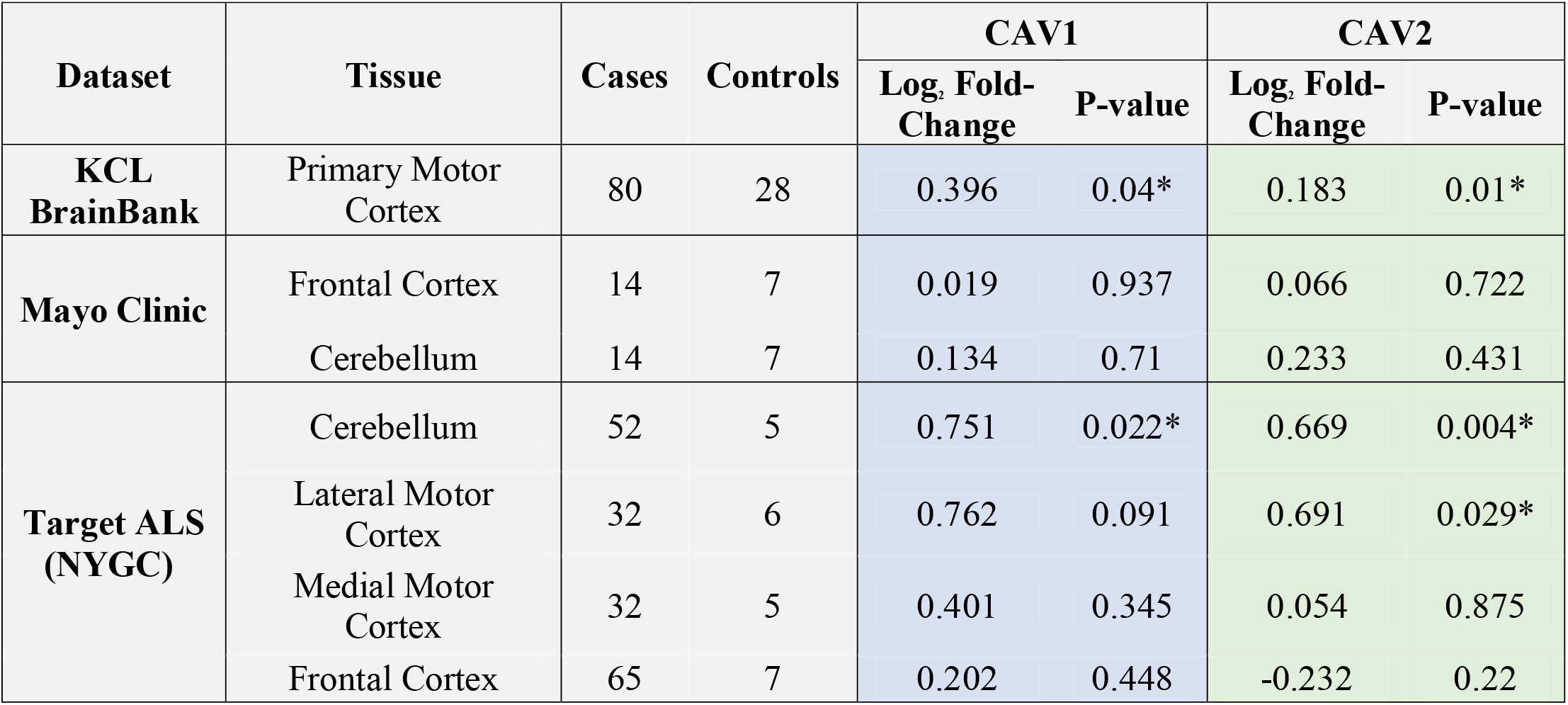
outlines the differential expression (Log_2_-fold change) for CAV1 (blue) and CAV2 (green) across brain tissues and datasets. *p < 0.05

| Dataset | Tissue | Cases | Controls | CAV1 |  | CAV2 |  |
| --- | --- | --- | --- | --- | --- | --- | --- |
|  |  |  |  | Log<sub>2</sub> Fold-Change | P-value | Log<sub>2</sub> Fold-Change | P-value |
| **KCL BrainBank** | Primary Motor Cortex | 80 | 28 | 0.396 | 0.04* | 0.183 | 0.01* |
| **Mayo Clinic** | Frontal Cortex | 14 | 7 | 0.019 | 0.937 | 0.066 | 0.722 |
|  | Cerebellum | 14 | 7 | 0.134 | 0.71 | 0.233 | 0.431 |
| **Target ALS (NYGC)** | Cerebellum | 52 | 5 | 0.751 | 0.022* | 0.669 | 0.004* |
|  | Lateral Motor Cortex | 32 | 6 | 0.762 | 0.091 | 0.691 | 0.029* |
|  | Medial Motor Cortex | 32 | 5 | 0.401 | 0.345 | 0.054 | 0.875 |
|  | Frontal Cortex | 65 | 7 | 0.202 | 0.448 | -0.232 | 0.22 |

**Figure 4.**
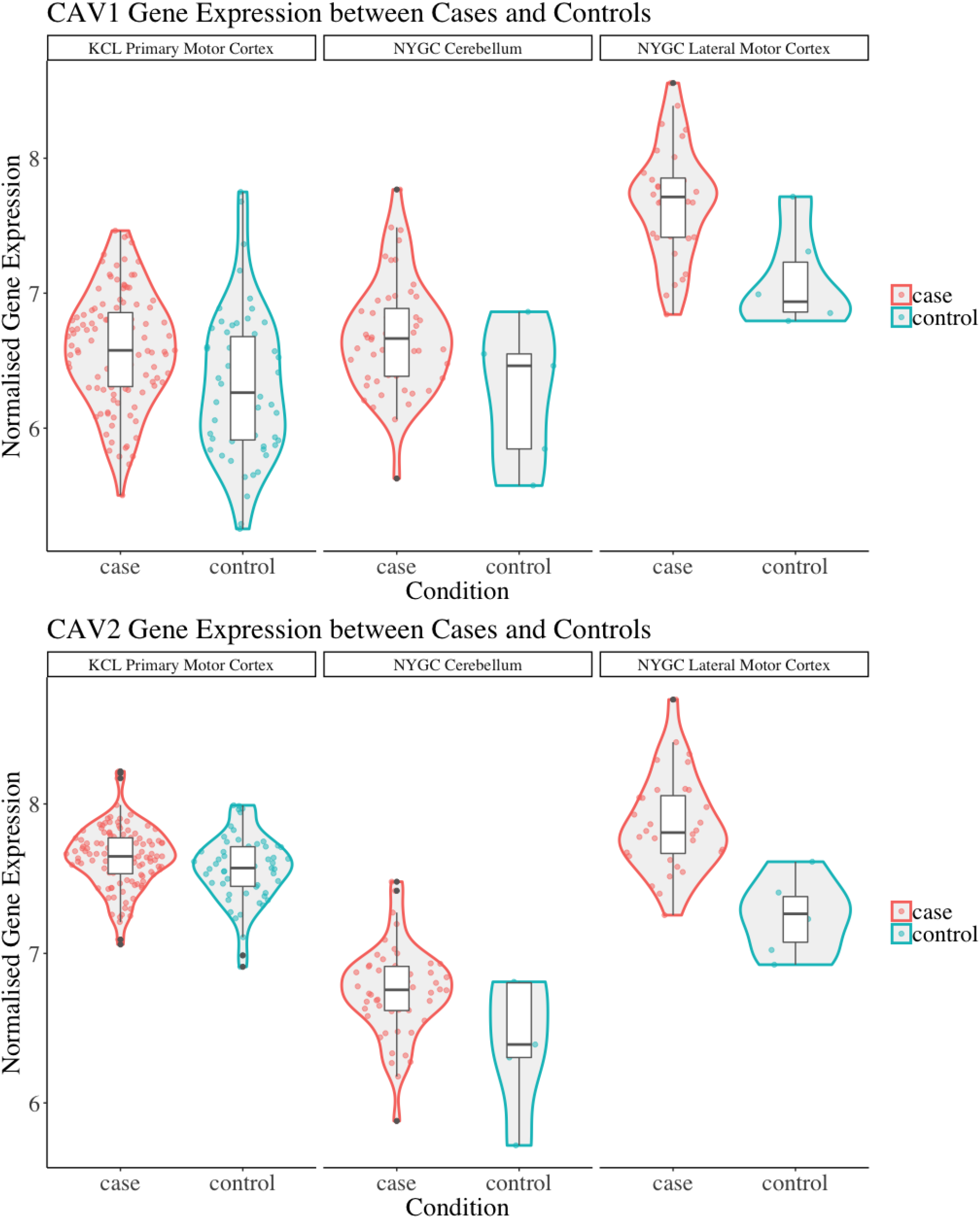
shows violin plots of significant gene expression for CAV1 and CAV2 between cases and controls. The X-axis indicates tissue/dataset combination and case/control status. The Y-axis is normalised gene expression. Coloured dots inside violin plots are jittered gene expressions for each sample. Boxplots inside each violin plot show gene expression for each category. Violin plot colour: Condition (case: red; control: blue). Note that CAV1 differential expression in the lateral motor cortex is significant only to p < 0.1. Violin plots for all analyses are available in Supplementary Figure 1.

Log_2_ fold-change for CAV1 and CAV2 were in a consistent direction across all datasets and tissues except for the CAV2 NYGC frontal cortex. For this reason, a stouffer meta-analysis was run for the motor cortex, frontal cortex, and cerebellum, the results of which are shown in *Table 2*. Two TargetALS NYGC tissue regions were available within the motor cortex, the lateral and medial motor cortex, of which only the lateral motor cortex reached statistical significance (Log2FC=0.691, *p*=0.029). These datasets were separately meta-analysed with the KCL Brainbank dataset. These analyses were statistically significant for both CAV1 and CAV2 genes, and all showed a large log2 fold-change over 2.

**Table 2.** Stouffer meta-analysis of differential expression data. *p < 0.05.

| Datasets | Tissue | Sample size 1 | Sample size 2 | CAV1 |  | CAV2 |  |
| --- | --- | --- | --- | --- | --- | --- | --- |
|  |  |  |  | Log<sub>2</sub> Fold-Change | P-value | Log<sub>2</sub> Fold-Change | P-value |
| KCL + NYGC (Lateral) | Motor Cortex | 108 | 38 | 2.499 | 0.012* | 3.155 | 0.002* |
| KCL + NYGC (Medial) | Motor Cortex | 108 | 37 | 2.249 | 0.025* | 2.488 | 0.013* |
| Mayo + NYGC | Frontal Cortex | 21 | 72 | 0.751 | 0.453 | -1.078 | 0.281 |
| Mayo + NYGC | Cerebellum | 21 | 57 | 2.280 | 0.023* | 2.969 | 0.003* |

### CAV1/2 expression is higher in iPSC-derived motor neurons from ALS patients

Bulk RNA-seq in *post-mortem* brain tissue has showed that expression of both *CAV1* and *CAV2* genes is higher in ALS patients compared to controls. Enhanced CAV1 expression has previously been associated with neuroprotection (24) and reduced CAV1 expression has been associated with risk for ALS (17). Therefore, the observed higher expression of CAV1 and CAV2 might represent a compensatory reaction to neurotoxicity. However, the bulk RNA-seq analysis does not allow us to determine which cell types are responsible for observed changes in *CAV1/2* expression. To address this, we analysed gene expression in iPSC-derived MNs from ALS patients (n = 55, https://www.answerals.org/) and neurologically normal controls (n = 15). Mean expression of both genes was higher in ALS patients compared to controls although this difference was not statistically significant (CAV1: mean ALS = 1.46 TPM, mean control = 1.3 TPM, *t* = 0.48, Log2FC = 0.1575, *p* = 0.31. CAV2: mean ALS = 1.67 TPM, mean control = 1.39 TPM, *t* = 1.43, Log2FC = 0.2647, *p*=0.08).

### Correlation analyses between CAV1/2 expression and phenotypic measures in AnswerALS

Using RNAseq from iPSC-derived MN, we examined the association between CAV1/2 expression and phenotypic measures. An outline of these results is shown in *Table 3*. Age of onset was quantified in days; there was no significant correlation between CAV1/2 expression and age of onset (Pearson correlation p>0.05). Survival was measured in days from date of onset to death and censored samples were not included because of the lack of longitudinal data; date of death was available for 27 ALS patients. Cox proportional hazards model was used to determine whether survival was significantly correlated with CAV1/2 expression. The first 10 principal components were used as covariates to control for population structure. Neither CAV1 (*p* = 0.96) nor CAV2 (*p* = 0.70) were significantly associated with survival in this cohort.

**Table 3.** Results of AnswerALS RNA-seq expression and phenotypic correlation analyses for CAV1 (blue) and CAV2 (green).

| Analysis | Test | CAV1 |  |  | CAV2 |  |  |
| --- | --- | --- | --- | --- | --- | --- | --- |
|  |  | Coefficient | t | P-value | Coefficient | t | P-value |
| iPSC Gene Expression | t-test | NA | 0.48 | 0.31 | NA | 1.43 | 0.08 |
| Age of Onset | Pearson Correlation | 0.13 | NA | 0.31 | -0.19 | NA | 0.39 |
| Survival | Cox Proportional Hazard | -0.02 | NA | 0.96 | 0.70 | NA | 0.21 |
| Disease Progression (ALSFRS Score) | Pearson's Correlation | -0.11 | -0.72 | 0.76 | -0.27 | -1.78 | 0.04 |

Next, we tested whether CAV1/2 expression was correlated with rate of change in ALSFRS, which is a measure of rate of disease progression. The ALSFRS was measured longitudinally between 2 and 10 times (with a median of four measurements). The delta-ALSFRS was calculated using linear regression based upon patient visit time and was available for 43 ALS patients. CAV2 expression but not CAV1 expression was negatively correlated with the rate of change of ALSFRS score (*figure 5*); iPSC-derived MN with higher CAV2 expression were derived from patients with a faster rate of decline in the ALSFRS (Pearson correlation *p* = 0.04, *t* = -1.78, *r* = -0.27). In view of our previous data, this could suggest that compensatory increase in CAV2 expression is highest in patients with more rapid disease progression. It is interesting that CAV1 has been previously associated with neuroprotection but was not significant in this test which may indicate opposing forces of compensatory upregulation with more aggressive disease and a therapeutic effect slowing disease progression.

**Figure 5.**
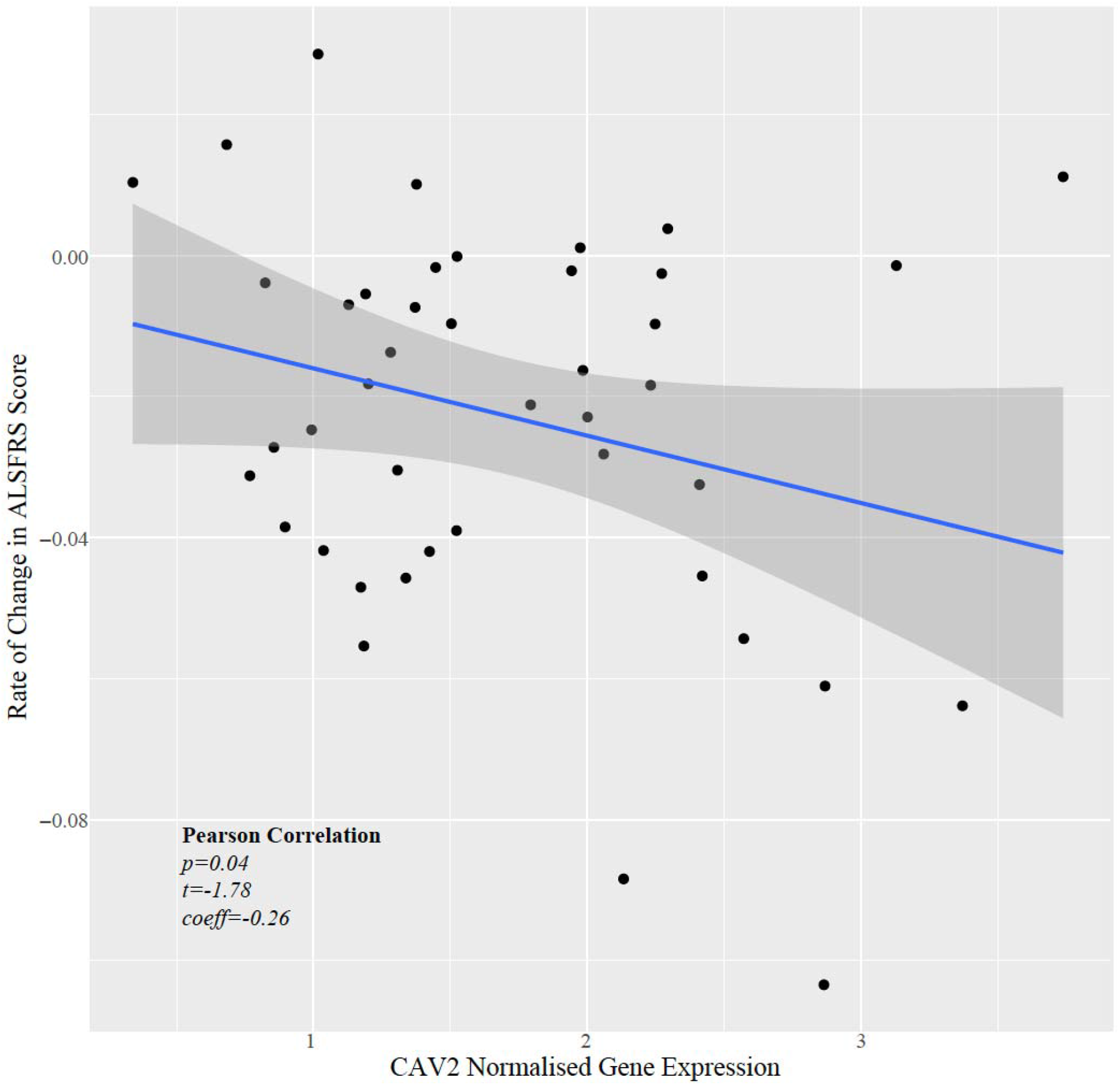
Scatter plot showing normalised CAV2 gene expression against rate of change in the ALSFRS.

**Figure 6.**
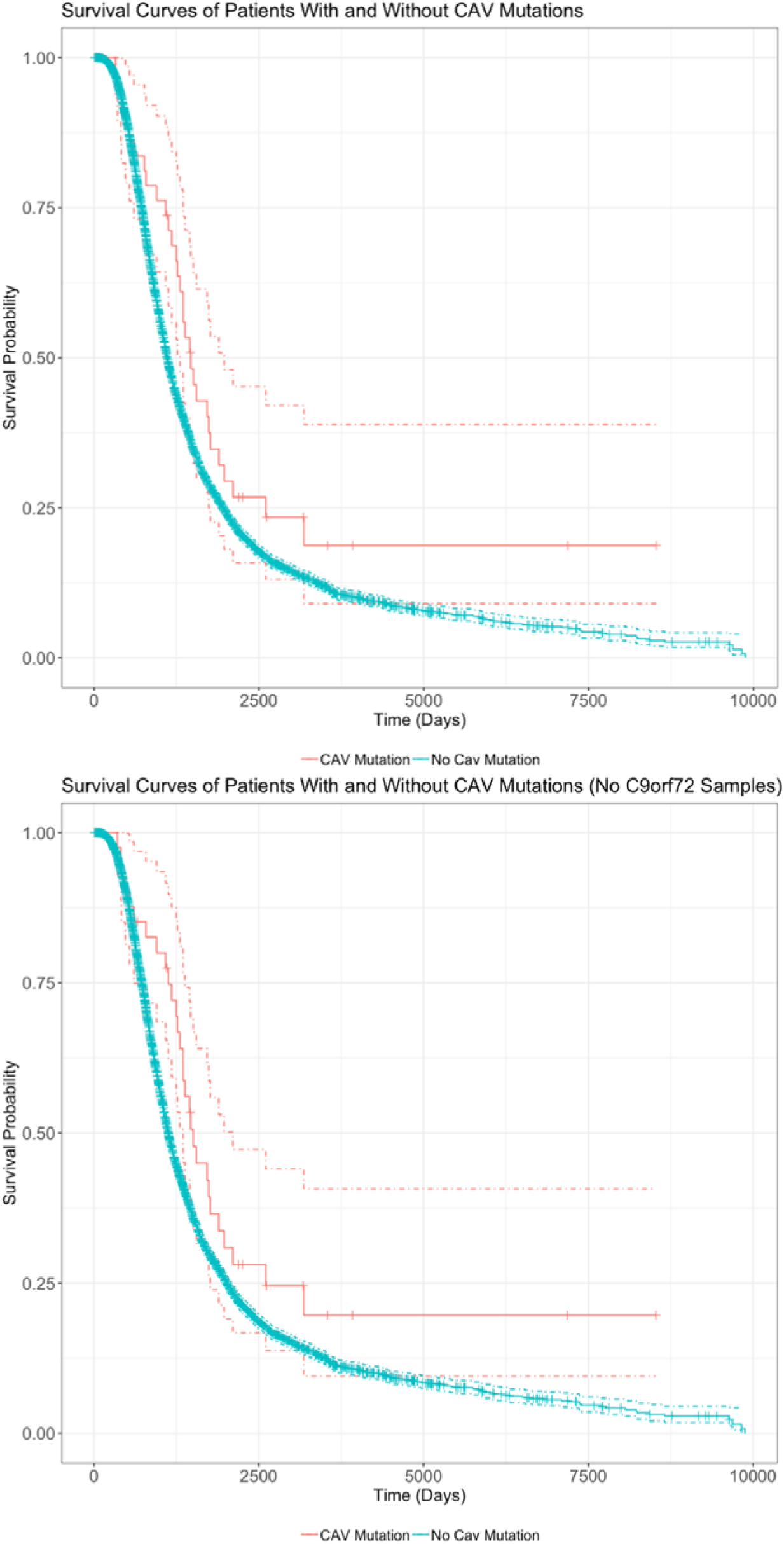
Survival Curves comparing survival of patients with vs without any CAV1/2 mutation. The top graph is based upon data from Analysis 1, inclusive of patients with C9orf72 repeat expansion. The bottom graph is from Analysis 2, with C9orf72 samples removed. Patients with CAV1/2 mutations have a longer survival time (C9orf72-inclusive analysis: median survival difference of 345 days. See table 5 for a full descriptive summary). Y-axis is the fraction of surviving sample. X-axis is time in days. Dashed lines indicate 95% confidence intervals. Orthogonal lines indicate death or censoring event. Graphs exclude 22 samples from patients surviving over 10000 days to improve scaling. Complete graphs available in Supplementary Figure 2.

### Survival Analyses in Project MinE

*Table 4* outlines the results from four of these survival analyses. In the first set of analyses (1-2) we tested the difference in survival of the patients carrying a mutation in the enhancer of either gene (CAV1/2) against non-carriers. The decision was made to combine CAV1 and CAV2 enhancer mutations due to their related biological function, co-expression, overlapping enhancers, and to maximise the statistical power. CAV1/2 mutations were significantly associated with longer survival (HR = 0.694, *p* = 0.043; HR = 0.674, *p* = 0.034). This was the case irrespective of whether *C9orf72* samples were included or removed.

**Table 4.** Breakdown of results across four survival analyses. The left side of the table displays the inclusion criteria of each analysis, and the right side displays the results. The first two column specifies whether samples with a C9orf72 mutation have been included (green tick) or excluded (red cross). CAV+ denotes the number of samples with CAV1/2 enhancer mutations, and CAV-indicates the sample size of those without CAV1/2 enhancer mutations. *p < 0.05; **p < 0.01; ***p < 0.001.

|  | C9orf72 | CAV Enhancer Mutations | CAV+ ALS | CAV- ALS |  | Hazard Ratio (95% CI) | Standard Error | p-value |
| --- | --- | --- | --- | --- | --- | --- | --- | --- |
| 1 | ✓ | CAV1 and CAV2 | 44 | 5943 | CAV | 0.694(0.487, 0.988) | 0.180 | 0.043* |
|  |  |  |  |  | Age of onset | 1.000 | 0.000 | < 0.001*** |
|  |  |  |  |  | Sex at birth | 1.075 (1.013, 1.141) | 0.031 | 0.018* |
| 2 | ✗ | CAV1 and CAV2 | 42 | 5589 | CAV | 0.674 (0.468, 0.971) | 0.186 | 0.034* |
|  |  |  |  |  | Age of onset | 1.000 | 0.000 | < 0.001*** |
|  |  |  |  |  | Sex at birth | 1.085 (1.020, 1.155) | 0.032 | 0.010** |
| 3 | ✗ | CAV1 | 33 | 5589 | CAV | 0.729 (0.484, 1.099) | 0.201 | 0.131 |
|  |  |  |  |  | Age of onset | 1.000 | 0.000 | < 0.001*** |
|  |  |  |  |  | Sex at birth | 1.084 (1.019, 1.154) | 0.032 | 0.011* |
| 4 | ✗ | CAV2 | 9 | 5589 | CAV | 0.523 (0.235, 1.164) | 0.409 | 0.112 |
|  |  |  |  |  | Age of onset | 1.000 | 0.000 | < 0.001*** |
|  |  |  |  |  | Sex at birth | 1.088 (1.022, 1.158) | 0.032 | 0.008** |

**Table 5.**
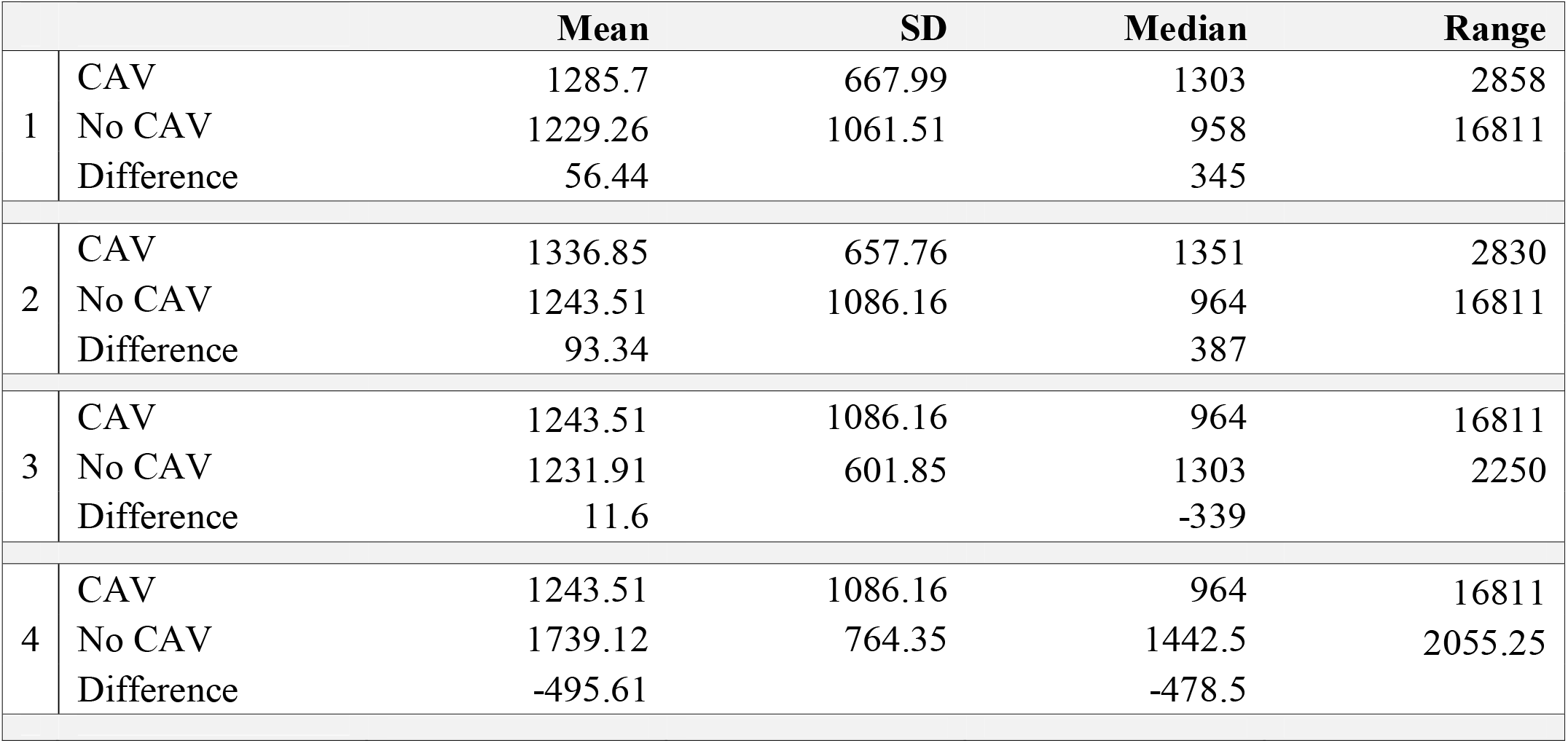
shows the mean, standard deviation, median, and range of survival in days for individuals with uncensored data. Analyses correspond to the rows in table 4. Analyses 1 and 2 are including and excluding patients with the C9orf72 repeat expansion, respectively. Analysis 3 and 4 are stratified by CAV1 and CAV2 enhancer mutation, and do not include C9orf72 mutations.

|  |  | Mean | SD | Median | Range |
| --- | --- | --- | --- | --- | --- |
| 1 | CAV | 1285.7 | 667.99 | 1303 | 2858 |
|  | No CAV | 1229.26 | 1061.51 | 958 | 16811 |
|  | Difference | 56.44 |  | 345 |  |
| 2 | CAV | 1336.85 | 657.76 | 1351 | 2830 |
|  | No CAV | 1243.51 | 1086.16 | 964 | 16811 |
|  | Difference | 93.34 |  | 387 |  |
| 3 | CAV | 1243.51 | 1086.16 | 964 | 16811 |
|  | No CAV | 1231.91 | 601.85 | 1303 | 2250 |
|  | Difference | 11.6 |  | -339 |  |
| 4 | CAV | 1243.51 | 1086.16 | 964 | 16811 |
|  | No CAV | 1739.12 | 764.35 | 1442.5 | 2055.25 |
|  | Difference | -495.61 |  | -478.5 |  |

The following analyses were then stratified by presence of CAV1 or CAV2 enhancer mutations. These analyses excluded *C9orf72* samples. Although not significant, the effects on survival of CAV1 and CAV2 enhancer mutations were similar and consistent with the analyses 1-2. This supports our initial choice to aggregate them to increase statistical power based on the hypothesis that mutations in the enhancers of both genes have a similar role in ALS.

### Age of Onset in Project MinE

Similarly to the survival analyses, each age of onset analysis was performed using differing inclusion criteria. *Table 6* displays the results for all age of onset analyses and their inclusion criteria. Cox Proportional hazards model was used for each analysis, setting the event status indicator to 1 (the event has occurred) for each sample. In parallel, a linear regression was performed using the same inclusion criteria as analysis 1. No analysis found any effect of CAV1/2 mutation on age of onset.

**Table 6.** The left side of the table describes the inclusion criteria and CAV+/ CAV-sample size. The right size shows results from age of onset analyses using Cox proportional hazards and linear regression models. ***p < 0.001.

|  | C9orf72 | CAV mutations | CAV+ ALS | CAV- ALS |  | Hazard Ratio (95% CI) | Standard Error | p-value |
| --- | --- | --- | --- | --- | --- | --- | --- | --- |
| 1 | ✓ | CAV1 and CAV2 | 44 | 5943 | CAV<br>Sex at birth | 1.034 (0.826, 1.296)<br>0.864 (0.820, 0.910) | 0.115<br>0.026 | 0.768<br>< 0.001*** |
| 2 | ✗ | CAV1 and CAV2 | 42 | 5589 | CAV<br>Sex at birth | 1.036 (0.823, 1.303)<br>0.859 (0.814, 0.906) | 0.117<br>0.027 | 0.764<br>< 0.001*** |
| 3 | ✗ | CAV1 | 33 | 5589 | CAV<br>Sex at birth | 1.078 (0.765, 1.518)<br>0.858 (0.813, 0.905) | 0.175<br>0.027 | 0.669<br>< 0.001*** |
| 4 | ✗ | CAV2 | 9 | 5589 | CAV<br>Sex at birth | 0.864 (0.449, 1.662)<br>0.857 (0.813, 0.905) | 0.334<br>0.027 | 0.662<br>< 0.001*** |
| Linear Regression |  |  |  |  |  | t- value | Standard Error | p-value |
| 5 | ✓ | CAV1 and CAV2 | 44 | 5943 | CAV<br>Sex at birth | -0.425<br>6.476 | 1.916<br>0.334 | 0.671<br>< 0.001*** |

## Discussion

We report increased expression of CAV1 and CAV2 in ALS cases when compared to controls using bulk RNA sequencing from *post-mortem* brain tissue samples. Statistically significant differential expression was found in the KCL Brainbank and Target ALS (NYGC) samples, but not in Mayo Clinic samples, although the direction of effect was consistent. Non-significant results may be due to a lack of power, as the sample size was substantially smaller in the Mayo Clinic samples than the other datasets. Additionally, meta-analyses revealed significant differences within the cerebellum and motor cortex for both CAV1 and CAV2 expression, but not the frontal cortex. One possible interpretation is that overexpression of CAV1/2 genes increases ALS risk. However, this is inconsistent with evidence that CAV1/2 expression is protective in ALS (17,23) and more generally promotes neuronal growth and improves motor function (38,39). An alternative interpretation consistent with previous literature is that the gene upregulation is indicative of a compensatory mechanism; CAV1/2 expression is increased as a response to ALS pathology, which affords greater protection.

Survival analyses showed that among ALS patients, carriers of CAV1/2 enhancer mutations had longer survival compared to non-carriers, with a median survival difference of 345 days in the Project MinE dataset. No correlation was demonstrated between gene expression and survival in the AnswerALS iPSC-derived MNs, although this analysis was limited by the small sample size. We observed a negative correlation between CAV2 expression and rate of change in the ALSFRS in the iPSC-derived MNs. Given the seemingly protective role of CAV1/2, it was expected that mutations in CAV1/2 enhancers, which purportedly decrease CAV1/2 expression, would in turn reduce survival. We consider two possible explanations for observing the opposite outcome. CAV1/2 enhancer mutations exist in non-coding regions and have unknown impact on gene expression. Cooper-Knock and colleagues (17) ran an expression analysis using a single CAV1/2 enhancer mutation (chr7:116222625:T>C), finding an association with reduced CAV1/2 expression in patient-derived neuronal cells. However, this is not sufficient evidence to conclude the global effect of CAV1/2 mutations on expression, as enhancer mutations may also increase gene expression (40,41). The effects of other variants on gene expression may account for the increased survival duration that we observed. Further investigation into the of CAV1/2 enhancer mutations on gene expression would be beneficial to build evidence for or against this interpretation.

An alternative hypothesis is that patients with CAV1/2 mutations represent a subset of ALS patients with a less aggressive phenotype. In this framework, CAV1/2 enhancer mutations reduce CAV1/2 expression, leading to dysfunctional neuronal signalling and accelerated neurodegeneration. However, the dysfunction associated with CAV1/2 is on average less severe than non-CAV-related ALS phenotypes, leading to the longer survival time found in our analyses. It is more likely that rare variants occurring within enhancer regions are deleterious, leading to reduced function of the enhancer and therefore reduced expression, than to improve function and increase CAV1/2 expression. This prior expectation makes this interpretation more biologically plausible.

Whether or not CAV1/2 enhancer mutations increase or decrease CAV1/2 gene expression, both align to the ‘compensatory model’ of CAV1/2 overexpression in ALS patients. If CAV1/2 are neuroprotective and are up regulated to compensate for ALS pathology, CAV1/2 enhancer mutations which increase expression simply boost this effect, leading to increased survival. If these mutations decrease expression and subsequently increase neurodegeneration, the ‘increased survival’ we observe among patients with CAV1/2 enhancer mutations may be explained by CAV-mediated ALS being on average less severe than non-CAV ALS.

Individuals with CAV1/2 mutations represent a small but relevant proportion of ALS patients (∼0.7%). Our results add to an increasing body of evidence linking CAV1 and CAV2 genes to ALS, help to elucidate the role of their enhancer mutations and gene expression in ALS, and support the positioning of CAV1/2 genes as potential targets for the development of treatment. However, further research into the functional effect of CAV1/2 mutations is needed to clarify their role in the pathogenesis of ALS.

## Data Availability

All data produced in the present study are available upon reasonable request to the authors.

https://www.ncbi.nlm.nih.gov/geo/query/acc.cgi?acc=GSE116622

https://www.ncbi.nlm.nih.gov/geo/query/acc.cgi?acc=GSE124439

https://www.ncbi.nlm.nih.gov/geo/query/acc.cgi?acc=GSE67196

https://www.projectmine.com/research/data-sharing/

https://dataportal.answerals.org/data-search

## Funding and Acknowledgments

We would like to acknowledge funding from the following funders: UK Research and Innovation; Medical Research Council; South London and Maudsley NHS Foundation Trust; MND Scotland; Motor Neurone Disease Association; National Institute for Health Research; Spastic Paraplegia Foundation; Rosetrees Trust; Darby Rimmer MND Foundation. Funding for open access charge: UKRI. B.N.A acknowledges funding from an NIHR pre-doctoral fellowship (NIHR301067). A.I is funded by the Motor Neurone Disease Association and South London and Maudsley NHS Foundation Trust. J.C.K. is supported by a Wellcome Trust fellowship (216596/Z/19/Z). AAK is funded by ALS Association Milton Safenowitz Research Fellowship (grant number22-PDF-609.DOI :10.52546/pc.gr.150909.), The Motor Neurone Disease Association (MNDA) Fellowship (Al Khleifat/Oct21/975-799), The Darby Rimmer Foundation, and The NIHR Maudsley Biomedical Research Centre. This is an EU Joint Programme-Neurodegenerative Disease Research (JPND) project. The project is supported through the following funding organizations under the aegis of JPND-http://www.neurodegenerationresearch.eu/ (United Kingdom, Medical Research Council MR/L501529/1 to A.A.-C., principal investigator [PI] and MR/R024804/1 to A.A.-C., PI]; Economic and Social Research Council ES/L008238/1 to A.A.-C. [co-PI]) and through the Motor Neurone Disease Association. This study represents independent research partly funded by the National Institute for Health Research (NIHR) Biomedical Research Centre at South London and Maudsley NHS Foundation Trust and King’s College London. The work leading up to this publication was funded by the European Community’s Horizon 2020 Programme (H2020-PHC-2014-two-stage; grant 633413). We acknowledge use of the research computing facility at King’s College London, Rosalind (https://rosalind.kcl.ac.uk), which is delivered in partnership with the National Institute for Health Research (NIHR) Biomedical Research Centres at South London & Maudsley and Guy’s & St. Thomas’ NHS Foundation Trusts and part-funded by capital equipment grants from the Maudsley Charity (award 980) and Guy’s and St Thomas’ Charity (TR130505).The views expressed are those of the author(s) and not necessarily those of the NHS, the NIHR, King’s College London, or the Department of Health and Social Care.

## References

1. Al-Chalabi A, Hardiman O. The epidemiology of ALS: a conspiracy of genes, environment and time. Nat Rev Neurol. 2013;9(11):617–28.

2. Lomen-Hoerth C, Murphy J, Langmore S, Kramer JH, Olney RK, Miller B. Are amyotrophic lateral sclerosis patients cognitively normal? Neurology. 2003;60(7):1094–7.

3. Strong MJ, Abrahams S, Goldstein LH, Woolley S, Mclaughlin P, Snowden J, et al. Amyotrophic lateral sclerosis-frontotemporal spectrum disorder (ALS-FTSD): Revised diagnostic criteria. Amyotroph lateral Scler Front Degener. 2017;18(3–4):153–74.

4. Conlon EG, Fagegaltier D, Agius P, Davis-Porada J, Gregory J, Hubbard I, et al. Unexpected similarities between C9ORF72 and sporadic forms of ALS/FTD suggest a common disease mechanism. Elife. 2018 Jul 13;7:e37754.

5. Phukan J, Pender NP, Hardiman O. Cognitive impairment in amyotrophic lateral sclerosis. Lancet Neurol. 2007 Nov;6(11):994–1003.

6. Al-Chalabi A, Fang F, Hanby MF, Leigh PN, Shaw CE, Ye W, et al. An estimate of amyotrophic lateral sclerosis heritability using twin data. J Neurol Neurosurg Psychiatry. 2010;81(12):1324–6.

7. Zou Z, Zhou Z, Che C, Liu C, … RH-J of N, 2017 U. Genetic epidemiology of amyotrophic lateral sclerosis: a systematic review and meta-analysis. jnnp.bmj.com. 2017;88(7):540–9.

8. Turner MR, Al-Chalabi A, Chio A, Hardiman O, Kiernan MC, Rohrer JD, et al. Genetic screening in sporadic ALS and FTD. J Neurol Neurosurg Psychiatry. 2017;88(12):1042–4.

9. Van Rheenen W, Shatunov A, Dekker AM, McLaughlin RL, Diekstra FP, Pulit SL, et al. Genome-wide association analyses identify new risk variants and the genetic architecture of amyotrophic lateral sclerosis. Nat Genet. 2016;48(9):1043.

10. Ryan M, Heverin M, McLaughlin RL, Hardiman O. Lifetime risk and heritability of amyotrophic lateral sclerosis. JAMA Neurol. 2019;76(11):1367–74.

11. Trabjerg BB, Garton FC, van Rheenen W, Fang F, Henderson RD, Mortensen PB, et al. ALS in Danish Registries: Heritability and links to psychiatric and cardiovascular disorders. Neurol Genet. 2020;6(2).

12. Brown RH, Al-Chalabi A. Endogenous retroviruses in ALS: A reawakening? Sci Transl Med. 2015;7(307):307fs40.

13. Braems E, Swinnen B, Van Den Bosch L. C9orf72 loss-of-function: a trivial, stand-alone or additive mechanism in C9 ALS/FTD? Acta Neuropathol. 2020;140(5):625–43.

14. Iacoangeli A, Al Khleifat A, Jones AR, Sproviero W, Shatunov A, Opie-Martin S, et al. C9orf72 intermediate expansions of 24–30 repeats are associated with ALS. Acta Neuropathol Commun. 2019;7(1):1–7.

15. Jones A, Iacoangeli A, Adey B, Reports HB-S, 2021 U. A HML6 endogenous retrovirus on chromosome 3 is upregulated in amyotrophic lateral sclerosis motor cortex. Sci Rep. 2021;11(1):1–10.

16. Young AI. Solving the missing heritability problem. PLoS Genet. 2019;15(6):e1008222.

17. Cooper-Knock J, Zhang S, Kenna KP, Moll T, Franklin JP, Allen S, et al. Rare variant burden analysis within enhancers identifies CAV1 as an ALS risk gene. Cell Rep. 2020;33(9):108456.

18. Theunissen F, Flynn LL, Anderton RS, Mastaglia F, Pytte J, Jiang L, et al. Structural variants may be a source of missing heritability in sALS. Front Neurosci. 2020;14:47.

19. Igarashi M, Honda A, Kawasaki A, Nozumi M. Neuronal Signaling Involved in Neuronal Polarization and Growth: Lipid Rafts and Phosphorylation. Vol. 13, Frontiers in Molecular Neuroscience. 2020. p. 150.

20. Yue K-Y, Zhang P-R, Zheng M-H, Cao X-L, Cao Y, Zhang Y-Z, et al. Neurons can upregulate Cav-1 to increase intake of endothelial cells-derived extracellular vesicles that attenuate apoptosis via miR-1290. Cell Death Dis. 2019;10(12):1–11.

21. Tang W, Li Y, Li Y, Wang Q. Caveolin-1, a novel player in cognitive decline. Neurosci Biobehav Rev. 2021;129:95–106.

22. Mandyam CD, Schilling JM, Cui W, Egawa J, Niesman IR, Kellerhals SE, et al. Neuron-targeted caveolin-1 improves molecular signaling, plasticity, and behavior dependent on the hippocampus in adult and aged mice. Biol Psychiatry. 2017;81(2):101–10.

23. Head BP, Hu Y, Finley JC, Saldana MD, Bonds JA, Miyanohara A, et al. Neuron-targeted caveolin-1 protein enhances signaling and promotes arborization of primary neurons. J Biol Chem. 2011;286(38):33310–21.

24. Sawada A, Wang S, Jian M, Leem J, Wackerbarth J, Egawa J, et al. Neuron□targeted caveolin□1 improves neuromuscular function and extends survival in SOD1G93A mice. FASEB J. 2019;33(6):7545–54.

25. Ichinomiya T, Wang S, Patel H, Tadokoro T, Marsala M, Head B. Subpial Gene Delivery of synapsin□promoted Caveolin□1 Prolongs Survival in hSODG93A mice Model of ALS. FASEB J. 35.

26. Head BP, Peart JN, Panneerselvam M, Yokoyama T, Pearn ML, Niesman IR, et al. Loss of caveolin-1 accelerates neurodegeneration and aging. PLoS One. 2010;5(12):e15697.

27. Iacoangeli A, Fogh I, Selvackadunco S, Topp SD, Shatunov A, van Rheenen W, et al. SCFD1 expression quantitative trait loci in amyotrophic lateral sclerosis are differentially expressed. Brain Commun. 2021;3(4):fcab236.

28. Smith L, Cupid BC, Dickie BGM, Al-Chalabi A, Morrison KE, Shaw CE, et al. Establishing the UK DNA Bank for motor neuron disease (MND). BMC Genet. 2015;16(1):1–8.

29. Zhang S, Cooper-Knock J, Weimer AK, Shi M, Moll T, Marshall JNG, et al. Genomewide identification of the genetic basis of amyotrophic lateral sclerosis. Neuron. 2022;110(6):992–1008.

30. Project MinE: study design and pilot analyses of a large-scale whole-genome sequencing study in amyotrophic lateral sclerosis. Eur J Hum Genet. 2018;26(10):1537–46.

31. Van Rheenen W, Van Der Spek RAA, Bakker MK, Van Vugt JJFA, Hop PJ, Zwamborn RAJ, et al. Common and rare variant association analyses in amyotrophic lateral sclerosis identify 15 risk loci with distinct genetic architectures and neuron-specific biology. Nat Genet. 2021;53(12):1636–48.

32. Baxi EG, Thompson T, Li J, Kaye JA, Lim RG, Wu J, et al. Answer ALS, a largescale resource for sporadic and familial ALS combining clinical and multi-omics data from induced pluripotent cell lines. Nat Neurosci. 2022;25(2):226–37.

33. Tam OH, Rozhkov N V, Shaw R, Kim D, Hubbard I, Fennessey S, et al. Postmortem Cortex Samples Identify Distinct Molecular Subtypes of ALS: Retrotransposon Activation, Oxidative Stress, and Activated Glia. Cell Press. 2019;

34. Prudencio M, Belzil V V., Batra R, Ross CA, Gendron TF, Pregent LJ, et al. Distinct brain transcriptome profiles in C9orf72-associated and sporadic ALS. Nat Neurosci. 2015 Aug 30;18(8):1175–82.

35. Stouffer SA, Suchman EA, DeVinney LC, Star SA, Williams Jr RM. The american soldier: Adjustment during army life.(studies in social psychology in world war ii), vol. 1. 1949;

36. Al-Chalabi A, Hardiman O, Kiernan MC, Chiò A, Rix-Brooks B, van den Berg LH. Amyotrophic lateral sclerosis: moving towards a new classification system. Lancet Neurol. 2016;15(11):1182–94.

37. Al-Chalabi A, Van Den Berg LH, Veldink J. Gene discovery in amyotrophic lateral sclerosis: implications for clinical management. Nat Rev Neurol. 2017;13(2):96.

38. Egawa J, Schilling JM, Cui W, Posadas E, Sawada A, Alas B, et al. Neuron□specific caveolin□1 overexpression improves motor function and preserves memory in mice subjected to brain trauma. FASEB J. 2017;31(8):3403–11.

39. Egawa J, Zemljic-Harpf A, Mandyam CD, Niesman IR, Lysenko L V, Kleschevnikov AM, et al. Neuron-targeted caveolin-1 promotes ultrastructural and functional hippocampal synaptic plasticity. Cereb Cortex. 2018;28(9):3255–66.

40. Sur I, Taipale J. The role of enhancers in cancer. Nat Rev Cancer. 2016;16(8):483–93.

41. Corradin O, Scacheri PC. Enhancer variants: evaluating functions in common disease. Genome Med. 2014;6(10):1–14.

