## Supplementary Figures for "Large-scale Analyses of CAV1 and CAV2 Suggest Their Expression is Higher in Post-mortem ALS Brain Tissue and Affects Survival"

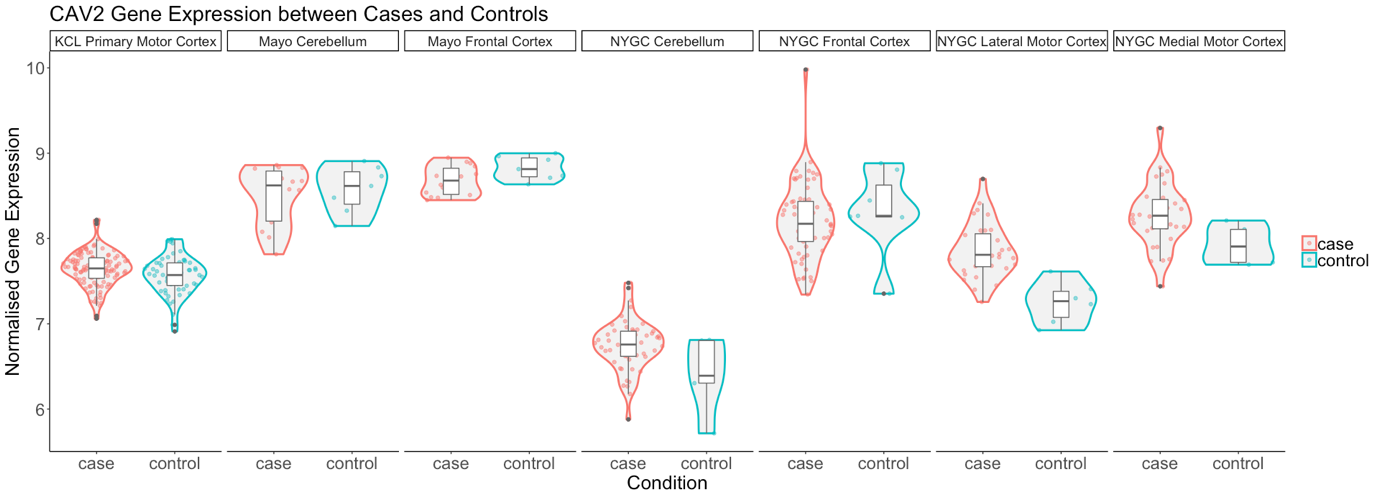

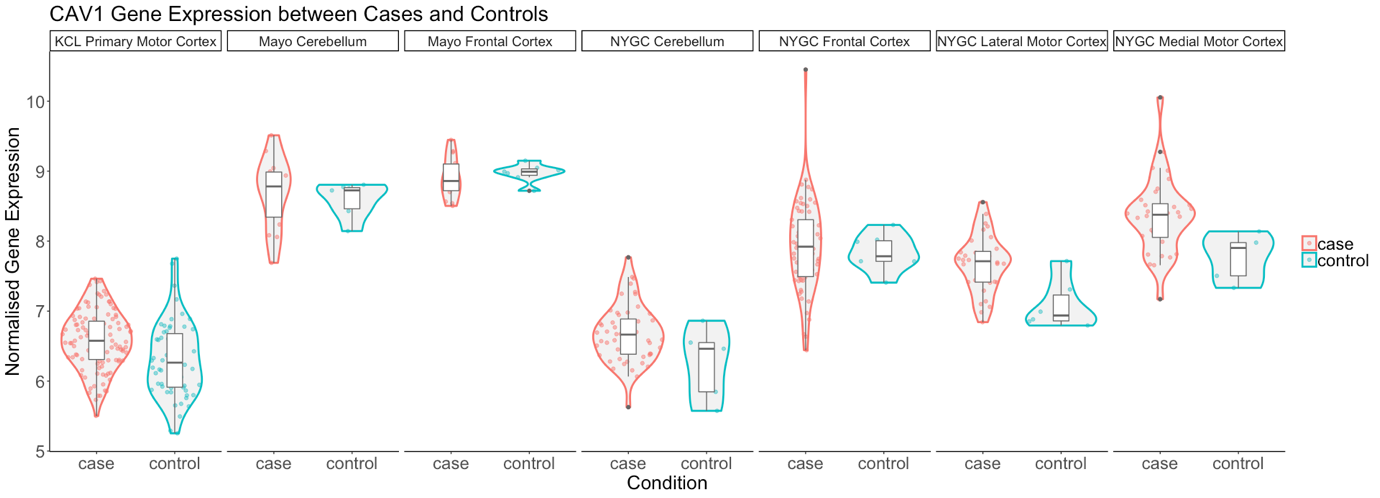

*Supplementary Figure 1. Violin plots of gene expression for CAV1 and CAV2 between cases and controls. The X-axis indicates tissue/dataset combination and case/control status. The Y-axis is normalised gene expression. Coloured dots inside violin plots are jittered gene expressions for each sample. Boxplots inside each violin plot show gene expression for each category. Violin plot colour: Condition (case: red; control: blue).*

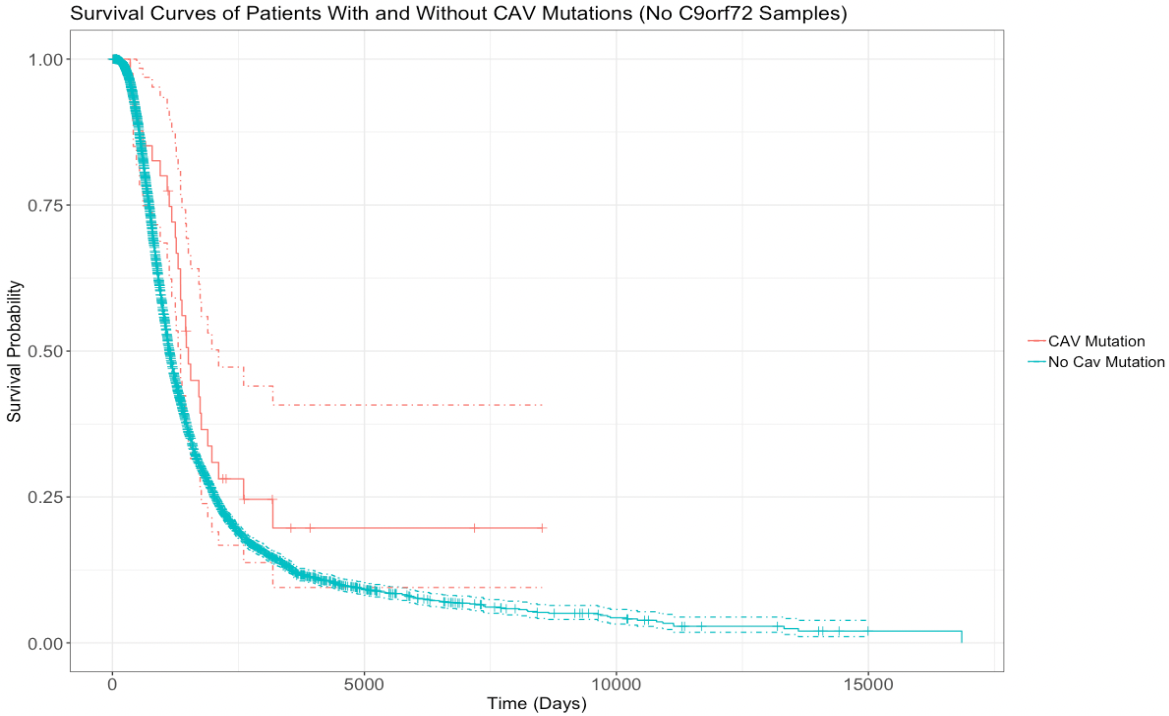

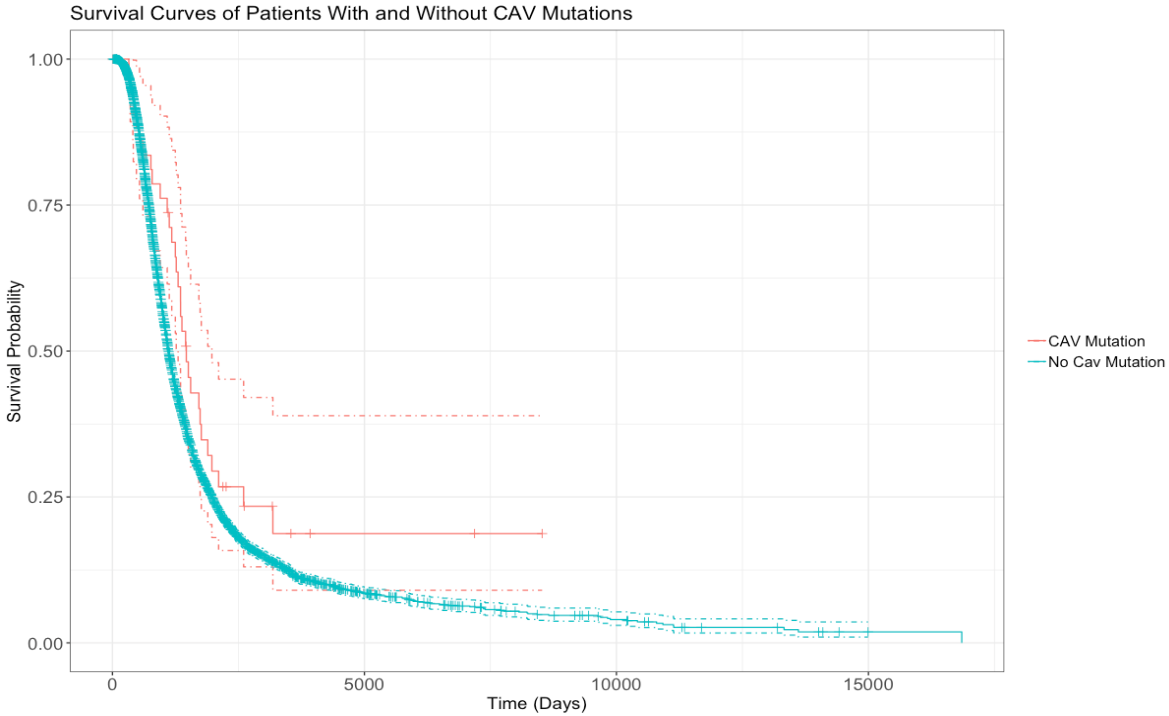

Supplementary Figure 2. Uncropped survival curves comparing survival of patients with vs without any CAV1/2 mutation. The top graph is based upon data from Analysis 1, inclusive of patients with C9orf72 repeat expansion. The bottom graph is from Analysis 2, with C9orf72 samples removed. Patients with CAV1/2 mutations have a longer survival time (C9orf72-inclusive analysis: median survival difference of 345 days. See table 5 for a full descriptive summary). Y-axis is the fraction of surviving sample. X-axis is time in days. Dashed lines indicate 95% confidence intervals. Orthogonal lines indicate death or censoring event.
